# CMR-Derived Myocardial Strain May Reflect Advanced Graft Injury Rather Than Acute Rejection After Heart Transplantation

**DOI:** 10.64898/2026.08.10.26360075

**Authors:** Marko Taipale, Markku Pentikäinen, Laura Martelius, Aino Mutka, Soili Kytölä, Matti Kankainen, Juha I. Peltonen, Simo Syrjälä, Arttu Lahtiharju, Jyri Lommi, Timo Jahnukainen, Karl Lemström, Tiina Ojala

**Author notes:** **Address for Correspondence**, Dr Marko Taipale, New Children’s Hospital, Pediatric Research Center, Pediatric Cardiology, PO Box 281, FI-00029 HUS, Stenbäckinkatu 11, Helsinki, Finland. equal contribution (Karl Lemström, Tiina Ojala).

## Abstract

**Aims:** Cardiac magnetic resonance imaging (CMR) T1 and T2 mapping accurately detect acute heart transplant rejection, but the diagnostic value of CMR-derived strain imaging remains uncertain, particularly for right ventricular strain. Data incorporating donor-derived cell-free DNA (dd-cfDNA) into a composite reference standard are limited. We evaluated the diagnostic accuracy of CMR-derived left and right ventricular strain and ejection fraction for detecting acute rejection in pediatric and adult heart transplant recipients.

**Methods and results:** Blinded analysis of 1.5T CMR studies was performed in pediatric and adult heart transplant recipients 1–24 months post-transplant and during five additional rejection episodes 3–14 years post-transplant. Left and right ventricular strain and ejection fraction were quantified using semi-automated post-processing. Acute rejection was defined using a composite reference standard comprising endomyocardial biopsy (EMB), clinical assessment, and dd-cfDNA. Diagnostic performance was assessed using receiver operating characteristic analysis. Among 214 CMR studies in 58 patients, 13 cases of acute rejection were identified. Diagnostic performance for detecting acute rejection was highest for pediatric right ventricular longitudinal strain (AUC 0.782, 95% CI 0.565–0.999), although imprecise due to data heterogeneity. All other cardiac functional parameters demonstrated limited discrimination in both pediatric and adult patients (AUC 0.536– 0.739). Models based on individual rejection indicators (EMB, clinical assessment, and dd-cfDNA) also showed poor diagnostic accuracy.

**Conclusion:** CMR-derived strain and ejection fraction showed limited diagnostic value for detecting acute rejection. Routine strain analysis therefore appears to have limited utility for rejection surveillance, with abnormalities primarily reflecting more advanced graft injury.

**Clinical Trial Registration:** ClinicalTrials.gov Identifier: NCT04311346

## Introduction

Although median graft survival after heart transplantation has improved, acute rejection remains a major clinical challenge. Within the first year post-transplant, it occurs in 13% of patients and accounts for 15% of deaths in children and 8% in adults.[1,2] While endomyocardial biopsy (EMB) has been the historical gold standard for rejection detection, it is not without limitations, including its invasiveness, potential for sampling error, and substantial inter-reporter variability.[3]

Cardiac magnetic resonance imaging (CMR) T1 and T2 mapping have shown high diagnostic accuracy for detecting acute rejection.[4–6] In contrast, the role of CMR-derived strain imaging in identifying early changes in myocardial mechanics remains less well defined, and studies incorporating the widely used biomarker donor-derived cell-free DNA (dd-cfDNA) as part of a composite reference standard are lacking.

Cardiac strain quantifies myocardial deformation, capturing left ventricular global longitudinal strain (GLS), global circumferential strain (GCS), and right ventricular longitudinal strain (RVLS), as measures of myocardial shortening during systole. In particular, GLS is generally more sensitive than left ventricular ejection fraction (LVEF) for detecting subtle or early systolic dysfunction.[7] However, as GLS is predominantly determined by longitudinally oriented myofibers in the left ventricle subendocardium, whereas rejection-related myocardial injury may be diffuse throughout the myocardium, GLS may not provide superior sensitivity for detecting acute rejection.

In heart transplant recipients, GLS, GCS, and RVLS are typically impaired compared with those in healthy controls and tend to improve over time after transplantation.[8–10] CMR studies have generally demonstrated reduced GLS and GCS during acute rejection, although findings have not been entirely consistent across studies, and RVLS has not been reported.[9–13] GLS and RVLS have also been shown to be independently associated with mortality and major adverse cardiac events in retrospective cohorts.[14] In contrast, studies evaluating LVEF and right ventricular ejection fraction (RVEF) have reported conflicting findings regarding their ability to detect acute rejection.[10,12,13]

This study aimed to evaluate the diagnostic accuracy of CMR-derived left and right ventricular strain and ejection fraction for detecting acute rejection in pediatric and adult heart transplant recipients. For this study, a composite reference standard incorporating EMB, clinical assessment, and dd-cfDNA was used to provide a broader and more clinically relevant definition of rejection.

## Methods

This prospective blinded study (Trial NCT04311346) was carried out at the pediatric and adult transplant units of Helsinki University Hospital. The study protocol received approval from the local ethics committee (HUS; HUS/3341/2019). Post-transplant surveillance followed the study protocol, with CMR, EMB, and dd-cfDNA assessments scheduled at 1, 2, 3, 4, 5, 6, and 12 months, followed by an additional CMR scan at 24 months. At each assessment point, acute rejection was diagnosed when at least two of the following three criteria were fulfilled: (1) EMB demonstrating grade ≥2R acute cellular rejection or grade ≥1 antibody-mediated rejection; (2) clinical signs or echocardiographic findings (reduced EF or hypertrophy) suggestive of rejection; and (3) dd-cfDNA levels exceeding 0.20%. When one criterion was unavailable, the diagnosis was established if either of the remaining two criteria was positive. All EMB specimens were graded according to International Society for Heart and Lung Transplantation criteria by a single pathologist. Clinical rejection was defined by agreement of the majority among three cardiologists, and dd-cfDNA concentrations were quantified using droplet digital PCR, with exclusion of samples in situations known to cause false-positive results.

CMR examinations from pediatric and adult heart transplant recipients were blindly analyzed between 1 and 24 months after transplantation. In addition, five acute rejection episodes from pediatric recipients occurring 3–14 years post-transplant were included.

All CMR examinations were performed on 1.5-T MRI scanners, with children <18 years scanned using Ingenia (Philips Healthcare, Best, the Netherlands) and adults ≥20 years using AvantoFit (Siemens Healthineers, Erlangen, Germany). Patients aged 18–20 years were classified according to the scanner used for the study. The imaging protocol included 2D balanced steady-state free precession (bSSFP) cine sequences acquired in short- and long-axis views. Left ventricular systolic deformation was assessed from cine images using feature-tracking–derived GLS and GCS, while right ventricular deformation was assessed using RVLS, which included both the septum and free wall. Volumetric parameters included LVEF, RVEF, as well as indexed left ventricular stroke volume (LVSV/BSA) and indexed cardiac output (LVCO/BSA). All measurements were obtained using semiautomated post-processing in Medis Suite 4.0 (Medis Medical Imaging Systems, Leiden, the Netherlands), with manual adjustment of ventricular contours performed when necessary.

Receiver operating characteristic (ROC) analyses, accounting for the clustered data, and Youden index were conducted to identify the exploratory thresholds for detecting acute rejection using strain and volumetric parameters. Linear mixed-effects models with a random intercept for patient, accounting for repeated measurements, were used to obtain estimates of strain and volumetric data. In sensitivity analyses, estimates were adjusted for the natural logarithm of time after transplantation, ln(time), and donor age. ROC analyses for clustered data were conducted with R version 4.5.2 (R Core Team, Vienna, Austria), and all other analyses with SPSS version 29.0 (IBM Corp, Armonk, New York).

The full methodological approach has been presented in previous publications.[5,6]

## Results

Among 214 CMR examinations performed in 58 patients, 13 episodes of acute rejection were identified, comprising five episodes in four pediatric patients and eight episodes in five adult patients (Table 1 and Table 2). Significant differences between the no-rejection and rejection groups were observed only for pediatric RVLS and indexed LVSV (Table 3). For pediatric RVLS, the estimated difference was 7.7 units (95% CI, 1.1–14.4; P=0.026) and remained similar after adjustment for ln(time) and donor age (β=8.0, 95% CI, −0.1 to 16.2; P=0.054) (Table 4). For LVSV/BSA, the difference attenuated from −7.9 ml/m^2^ (95% CI, −15.0 to −0.7; P=0.033) to −3.7 ml/m^2^ (95% CI, −13.0 to 5.5; P=0.411) after adjustment. Diagnostic accuracy was moderate for pediatric RVLS (AUC 0.782, 95% CI 0.565–0.999) (Table 5). In contrast, all other cardiac functional parameters demonstrated limited discriminatory performance, with AUCs ranging from 0.536 to 0.739. Inadequate diagnostic accuracy was also observed in strain models relying on single rejection indicators (EMB, clinical signs, and dd-cfDNA) (Table 6).

**Table 1.** Baseline patient characteristics*.

|  | **Pediatric Transplants**<br>**n = 17** | **Adult Transplants**<br>**n = 41** |
|---|---|---|
| **Sex, male** | 7 (41%) | 33 (80%) |
| **Age at HTx (yrs)** | 12.3 (10.3-13.3) | 54.7 (45.2-61.7) |
| **BMI at HTx (kg/m2)** |  | 25.5 (22.5-27.7) |
| **ISO-BMI** | 19.7 (17.4-26.9) |  |
| **Donor age (yrs)** | 19 (14-23) | 38 (31-46) |
| **Ischemic time (min)** | 205 (146-251) | 186 (117-214) |
| **No. of scans per patient** | 1 (1-3) | 4 (3-6) |
| **CMR, mean time from HTx (mo)‡** | 20.2 (7.5-27.8) | (3.9-5.6) |

Values are median (interquartile range), or n (%) unless otherwise stated.
\*Reproduced from Taipale et al., Eur Heart J Cardiovasc Imaging (2026), https://doi.org/10.1093/ehjci/jeag080, CC BY 4.0. ‡ Includes CMR studies from later time points, which correspond to rejection cases in the pediatric group.
BMI, body mass index; CMR, cardiac magnetic resonance imaging; HTx, heart transplantation; ISO-BMI, age- and sex-adjusted body mass index.

**Table 2.** Individual rejection parameters.

|  | **Children** | **Adults** |
|---|---|---|
| **EMB** | n = 31 | n = 175 |
| **ACR grade 0R** | 22 (71%) | 107 (61%) |
| **ACR grade 1R** | 6 (19%) | 61 (35%) |
| **ACR grade 2-3R** | 3 (10%) | 7 (4%) |
| **pAMR 0** | 30 (97%) | 171 (98%) |
| **pAMR 1-3** | 1 (3%) | 4 (2%) |
| **Clinical assessment** | n = 33 | n = 181 |
| **No rejection** | 27 (82%) | 176 (97%) |
| **Rejection** | 6 (18%) | 5 (3%) |
| **dd-cfDNA** | n = 5 | n = 157 |
| **≤ 0.20%** | 5 (100%) | 144 (92%) |
| **> 0.20%** | 0 | 13 (8%) |

ACR, acute cellular rejection; dd-cfDNA, donor-derived cell-free DNA; EMB, endomyocardial biopsy; pAMR, pathologic antibody-mediated rejection

**Table 3.** Cardiac functional parameter estimates.

|  | **No rejection<br>(201 scans)** | **Rejection<br>(13 scans)** | **P value** |
|---|---|---|---|
| **Children** | 28 scans | 5 scans |  |
| **GLS (%)** | -23.0 (-26.1 to -19.9) | -20.2 (-25.8 to -14.5) | 0.365 |
| **GCS (%)** | -34.8 (-39.4 to -30.2) | -28.0 (-36.8 to -19.1) | 0.163 |
| **RVLS (%)** | -24.7 (-27.8 to -21.5) | -17.0 (-22.9 to -11.1) | **0.026** |
| **LVEF (%)** | 61.2 (55.7 to 66.7) | 50.5 (40.3 to 60.6) | 0.067 |
| **RVEF (%)** | 54.5 (49.8 to 59.2) | 45.7 (37.0 to 54.3) | 0.076 |
| **LVSV/BSA (ml/m2)** | 44.6 (41.3 to 47.9) | 36.7 (30.3 to 43.1) | **0.033** |
| **LVCO/BSA (l/min/m2)** | 3.8 (3.5 to 4.1) | 3.6 (3.0 to 4.2) | 0.531 |
| **Adults** | 173 scans | 8 scans |  |
| **GLS (%)** | -22.7 (-24.5 to -20.8) | -22.1 (-26.2 to -18.1) | 0.784 |
| **GCS (%)** | -30.4 (-32.7 to -28.1) | -29.2 (-33.9 to -24.4) | 0.579 |
| **RVLS (%)** | -25.4 (-27.4 to -23.4) | -23.0 (-27.4 to -18.6) | 0.256 |
| **LVEF (%)** | 56.4 (53.9 to 59.0) | 54.6 (50.0 to 59.2) | 0.379 |
| **RVEF (%)** | 55.0 (52.6 to 57.3) | 52.1 (47.6 to 56.7) | 0.174 |
| **LVSV/BSA (ml/m2)** | 43.7 (41.3 to 46.1) | 44.0 (39.2 to 48.8) | 0.885 |
| **LVCO/BSA (l/min/m2)** | 3.8 (3.6 to 4.0) | 3.6 (3.1 to 4.0) | 0.311 |

Values are linear mixed-effect model estimates (95% confidence interval), with p-values for group comparisons. In adult no-rejection cases, LVEF and RVEF were available in 172 cases, whereas LVSV/BSA and LVCO/BSA were available in 171 cases.
GCS, global circumferential strain; GLS, global longitudinal strain; LVCO/BSA, indexed left ventricular cardiac output; LVEF, left ventricular ejection fraction; LVSV/BSA, indexed left ventricular stroke volume; RVEF, right ventricular ejection fraction; RVLS, right ventricular longitudinal strain.

**Table 4.** Differences in cardiac functional parameters between acute rejection and no-rejection groups.

|  | **Unadjusted β<br>(95% CI)** | **P value** | **Adjusted for ln(time) and donor age β (95% CI)** | **P value** |
|---|---|---|---|---|
| **Children** |  |  |  |  |
| **GLS (%)** | 2.8 (-3.6 to 9.3) | 0.365 | 1.6 (-6.3 to 9.5) | 0.675 |
| **GCS (%)** | 6.9 (-3.1 to 16.8) | 0.163 | 8.5 (-3.8 to 20.6) | 0.163 |
| **RVLS (%)** | 7.7 (1.1 to 14.4) | 0.026 | 8.0 (-0.1 to 16.2) | 0.054 |
| **LVEF (%)** | -10.7 (-22.3 to 0.8) | 0.067 | -13.6 (-27.9 to 0.7) | 0.062 |
| **RVEF (%)** | -8.8 (-18.7 to 1.0) | 0.076 | -9.6 (-21.9 to 2.7) | 0.118 |
| **LVSV/BSA (ml/m2)** | -7.9 (-15.0 to -0.7) | 0.033 | -3.7 (-13.0 to 5.5) | 0.411 |
| **LVCO/BSA (l/min/m2)** | -0.2 (-0.9 to 0.5) | 0.531 | -0.1 (-1.0 to 0.8) | 0.825 |
| **Adults** |  |  |  |  |
| **GLS (%)** | 0.5 (-3.3 to 4.4) | 0.784 | 0.6 (-3.3 to 4.5) | 0.761 |
| **GCS (%)** | 1.2 (-3.2 to 5.6) | 0.579 | 1.7 (-2.7 to 6.0) | 0.442 |
| **RVLS (%)** | 2.4 (-1.8 to 6.6) | 0.256 | 2.5 (-1.7 to 6.7) | 0.245 |
| **LVEF (%)** | -1.8 (-5.9 to 2.3) | 0.379 | -2.5 (-6.4 to 1.4) | 0.204 |
| **RVEF (%)** | -2.8 (-6.9 to 1.3) | 0.174 | -3.3 (-7.3 to 0.8) | 0.117 |
| **LVSV/BSA (ml/m2)** | 0.3 (-4.1 to 4.7) | 0.885 | -0.2 (-4.5 to 4.2) | 0.945 |
| **LVCO/BSA (l/min/m2)** | -0.2 (-0.7 to 0.2) | 0.311 | -0.3 (-0.7 to 0.2) | 0.217 |

β = The estimate for difference between rejection and no rejection groups calculated using a linear mixed-effect model.
CI, confidence interval; GCS, global circumferential strain; GLS, global longitudinal strain; LVCO/BSA, indexed left ventricular cardiac output; LVEF, left ventricular ejection fraction; LVSV/BSA, indexed left ventricular stroke volume; RVEF, right ventricular ejection fraction; RVLS, right ventricular longitudinal strain.

**Table 5.** Diagnostic accuracy of cardiac functional parameters in detecting acute rejection.

|  | n | AUC (95% CI) | Exploratory cutoff* | Sensitivity (%) | Specificity (%) |
|---|---|---|---|---|---|
| **Children** |  |  |  |  |  |
| **GLS** | 33 | 0.682 (0.315-1.000) | ≥ -21 | 80 | 57.1 |
| **GCS** | 33 | 0.657 (0.376-0.938) | ≥ -33 | 80 | 57.1 |
| **RVLS** | 33 | **0.782 (0.565-0.999)** | ≥ -23 | 100 | 46.4 |
| **LVEF** | 33 | 0.696 (0.430-0.963) | ≤ 46 | 40 | 92.9 |
| **RVEF** | 33 | 0.643 (0.349-0.937) | ≤ 41 | 40 | 92.9 |
| **LVSV/BSA** | 33 | 0.739 (0.327-1.000) | ≤ 33 | 60 | 100 |
| **LVCO/BSA** | 33 | 0.564 (0.195-0.934) | ≤ 3.1 | 40 | 92.9 |
| **Adults** |  |  |  |  |  |
| **GLS** | 181 | 0.639 (0.341-0.937) | ≥ -11 | 37.5 | 98.3 |
| **GCS** | 181 | 0.600 (0.285-0.916) | ≥ -17 | 37.5 | 96.5 |
| **RVLS** | 181 | 0.633 (0.345-0.921) | ≥ -18 | 50 | 85 |
| **LVEF** | 179 | 0.654 (0.384-0.925) | ≤ 38 | 37.5 | 98.8 |
| **RVEF** | 179 | 0.615 (0.303-0.927) | ≤ 44 | 37.5 | 97.1 |
| **LVSV/BSA** | 178 | 0.575 (0.293-0.857) | ≤ 38 | 50 | 71.2 |
| **LVCO/BSA** | 178 | 0.536 (0.318-0.754) | ≤ 4.1 | 87.5 | 29.4 |

Optimal thresholds were identified using ROC analysis and Youden index.
\* GLS, GCS, and RVLS are expressed as percentages (%); LVSV/BSA as ml/m²; and LVCO/BSA as l/min/m².
AUC, area under the curve; CI, confidence interval; GCS, global circumferential strain; GLS, global longitudinal strain; LVCO/BSA, indexed left ventricular cardiac output; LVEF, left ventricular ejection fraction; LVSV/BSA, indexed left ventricular stroke volume; RVEF, right ventricular ejection fraction; RVLS, right ventricular longitudinal strain.

**Table 6.** Diagnostic accuracy of myocardial strain to detect acute rejection defined by EMB, dd-cfDNA, and clinical data.

|  | **n<br>(total/rej)** | **Exploratory<br>cutoff\*** | **Comparison** | **Sensitivity<br>(%)** | **Specificity<br>(%)** |
|---|---|---|---|---|---|
| **Children** |  |  |  |  |  |
| **GLS** | 31/3 | ≥ -21 % | EMB | 66.7 | 53.6 |
| **GLS** | 33/6 | ≥ -21 % | Clinical | 83.3 | 59.3 |
| **GCS** | 31/3 | ≥ -33 % | EMB | 100 | 57.1 |
| **GCS** | 33/6 | ≥ -33 % | Clinical | 83.3 | 59.3 |
| **RVLS** | 31/3 | ≥ -23 % | EMB | 100 | 42.9 |
| **RVLS** | 33/6 | ≥ -23 % | Clinical | 100 | 48.1 |
| **Adults** |  |  |  |  |  |
| **GLS** | 175/10 | ≥ -11 % | EMB | 10 | 97 |
| **GLS** | 157/13 | ≥ -11 % | dd-cfDNA | 7.7 | 98.6 |
| **GLS** | 181/5 | ≥ -11 % | Clinical | 40 | 97.7 |
| **GCS** | 175/10 | ≥ -17 % | EMB | 10 | 95.2 |
| **GCS** | 157/13 | ≥ -17 % | dd-cfDNA | 7.7 | 97.9 |
| **GCS** | 181/5 | ≥ -17 % | Clinical | 40 | 96 |
| **RVLS** | 175/10 | ≥ -18 % | EMB | 20 | 84.2 |
| **RVLS** | 157/13 | ≥ -18 % | dd-cfDNA | 15.4 | 84.7 |
| **RVLS** | 181/5 | ≥ -18 % | Clinical | 40 | 84.1 |

\* Cut-off values derived from the model using composite rejection criteria. Inadequate number of dd-cfDNA samples in children.
dd-cfDNA, donor-derived cell-free DNA; EMB, endomyocardial biopsy; GCS, global circumferential strain; GLS, global longitudinal strain; rej, rejected; RVLS, right ventricular longitudinal strain.

## Discussion

This prospective blinded study demonstrated that CMR-derived left and right ventricular strain imaging and ejection fraction have limited diagnostic accuracy for detecting acute rejection in both pediatric and adult heart transplant recipients. Notably, only RVLS in pediatric patients showed discriminatory performance to detect acute rejection.

In our cohort, the more severe rejection cases occurred in teenagers with reduced adherence to immunosuppressive therapy. Among the three pediatric patients with the most severe rejection, two had biopsy-proven ACR 3R, one of whom developed multiorgan failure. RVLS showed dynamic abnormalities—impairment during rejection and/or subsequent improvement during recovery—in all three cases, whereas similar changes in GLS and GCS were observed in two. In contrast, such severe presentations were not observed in adults. This may explain the observed reduction in RVLS and indexed LVSV only in pediatric rejection cases, and it is possible that a larger number of rejection episodes would have revealed additional abnormalities in other functional parameters.

In the study by Pouliopoulos et al., the authors report moderate to good performance of GCS (AUC 0.852) and GLS (AUC 0.742) for detecting histologically significant rejection (≥2R), with further improvement when strain is combined with T1 and T2 mapping.[11] The apparent discrepancy with our findings is more plausibly explained by differences in the definition of rejection and the biological stage of graft injury being captured.

In our study, acute rejection was defined using a composite reference standard incorporating EMB, clinical assessment, and dd-cfDNA. This approach strengthens the robustness of the reference standard and reflects real-world diagnostic complexity, and may capture inflammatory graft injury at an earlier stage. At this stage, tissue-level abnormalities detectable by T1 and T2 mapping, as well as elevations in dd-cfDNA, may already be present, whereas measurable changes in global myocardial strain may not yet have developed. Consistent with this, we observed significant elevations in T1 mapping (and, in pediatric patients, T2 mapping) during rejection, as well as associations between dd-cfDNA and mapping parameters.[5] Moreover, T1 mapping values showed positive correlations with strain and ejection fraction in univariable analysis, supporting a continuum from tissue-level injury to functional impairment. In contrast, studies focusing on histologically significant ≥2R rejection are more likely to capture later-stage disease with more pronounced functional impairment, in which strain abnormalities become more evident.

From this perspective, our findings and those of previous studies are complementary rather than contradictory. CMR mapping techniques and circulating biomarkers may be more sensitive to early inflammatory graft injury, whereas strain parameters likely reflect downstream myocardial dysfunction in more advanced rejection. Rather than serving as robust standalone markers of acute rejection, strain and ejection fraction may therefore be better interpreted as indicators of more established or clinically overt disease. In particular, RVLS should not be interpreted in isolation, as longitudinal strain may not fully capture right ventricular adaptation and global performance after transplantation.

Taken together with our previous work demonstrating excellent diagnostic accuracy of 16-segment T1 mapping (AUC 0.940–0.979) and pediatric T2 mapping (AUC 0.978) for detecting acute rejection, CMR-derived mapping and functional parameters may reflect a progressive spectrum of rejection-related myocardial injury.[6] We hypothesize that tissue characterization abnormalities may precede measurable changes in myocardial strain and ejection fraction during rejection. Future studies integrating tissue characterization, circulating biomarkers such as dd-cfDNA, and myocardial mechanics will be essential to establish a staged, multiparametric framework for non-invasive rejection surveillance.

This study has several limitations, including its single-center design, the absence of dd-cfDNA measurements at the 24-month time point, and the use of different imaging vendors for pediatric and adult cohorts. The relatively low number of rejection events and exclusion of the early high-risk post-transplant period reduce statistical precision. Furthermore, the lack of systematic examinations before, during, and after rejection episodes limited reliable longitudinal assessment of strain trajectories, while incomplete data on immunosuppressive treatment and comorbidities may have introduced residual confounding. Although rejection cases meeting only 1 of 2 criteria introduce heterogeneity into the rejection definition, most of these cases had multiple clinical signs of rejection.

## Conclusions

CMR-derived left and right ventricular strain and ejection fraction showed limited diagnostic accuracy for detecting acute rejection. Thus, routine CMR strain analysis appears to offer little utility for rejection surveillance, with strain abnormalities primarily reflecting more advanced graft injury.

## Abbreviations

AUC: (area under the curve
CI: (confidence interval
dd-cfDNA: (donor-derived cell-free DNA
EMB: (endomyocardial biopsy
GCS: (global circumferential strain
GLS: (global longitudinal strain
LVCO/BSA: (indexed left ventricular cardiac output
LVEF: (left ventricular ejection fraction
LVSV/BSA: (indexed left ventricular stroke volume
NPV: (negative predictive value
PPV: (positive predictive value
ROC: (receiver operator characteristics
RVEF: (right ventricular ejection fraction
RVLS: (right ventricular longitudinal strain).

## Data Availability

The data underlying this article cannot be shared publicly due to ethical reasons. The data is available for review by onsite visitors.

## Acknowledgements

The authors thank Tero Vahlberg for statistical analysis.

## Generative AI statement

The authors declared that generative AI was used in the creation of this manuscript. ChatGPT (OpenAI) was used for language editing only, and the authors take full responsibility for the final text and its content.

